# Do National Health Insurance Schemes Guarantee Financial Risk Protection in the drive towards Universal Health Coverage in West Africa? A Systematic Review of Observational Studies

**DOI:** 10.1101/2022.10.27.22281514

**Authors:** Sydney N.N.T. Odonkor, Ferdinand Koranteng, Martin Appiah-Danquah, Lorena Dini

## Abstract

**Background:** To facilitate the drive towards Universal Health Coverage (UHC) several countries in the West African subregion have over the last two decades adopted the system of National Health Insurance (NHI) to finance their health services. However, most of these countries continue to face challenges safeguarding the insured population against catastrophic health expenditure (CHE) and impoverishment due to health spending. The aim of this study is to describe the extent of financial risk protection among households enrolled under NHI schemes in West Africa and summarize potential learnings.

**Methods:** We conducted a systematic review of observational studies in accordance with the Preferred Reporting Items for Systematic Reviews and Meta-Analyses (PRISMA) guidelines. Studies published in English between 2005 and 2022 were searched for using keywords, synonyms and MeSH terms related to NHI, financial risk protection and UHC in all West African countries on the following electronic databases: PubMed/Medline, Web of Science and CINAHL via EBSCOhost and Embase via Ovid and Google Scholar. The quality of included studies was assessed using the Joanna Briggs Institute (JBI) critical appraisal checklist. Two independent reviewers assessed the studies for inclusion, extracted data and conducted quality assessment. We present the findings of the narrative synthesis consisting of thematic synthesis for qualitative data and a Synthesis Without Meta-analysis (SWiM) for quantitative data. The study protocol was published in PROSPERO under the ID CRD42022338574 on 28th June 2022.

**Results:** Of the 1,279 articles initially identified, nine were eligible for inclusion. These were cross-sectional studies (n=8) and retrospective cohort study (n=1) published between 2011 and 2021 in Ghana (n=8) and Nigeria (n=1). Two-thirds of the included studies reported that enrollment into the NHI showed a positive (protective) effect on CHE at different thresholds and one study showed a protective effect of NHI on impoverishment due to health spending.

However, almost all of the included studies (n=8) reported that a proportion of insured households still encountered CHE with one-third of them reporting more than 50% of insured households incurring CHE. Key determinants of CHE and impoverishment due to health spending reported consisted of income, employment and educational status of household members as well as household size, household health profile, gender of household head and distance of household from health facility.

**Discussion:** Households insured under NHI schemes in some West African countries (Ghana and Nigeria) are better protected against CHE and impoverishment due to health spending compared to uninsured households as evidenced in other studies. However, insured households continue to incur CHE and impoverishment due to health expenditure resulting from gaps identified in the current design of NHI schemes in these West African countries.

**Conclusion:** To protect insured households from the financial burden due to health spending and advance the drive towards UHC in West Africa, governments should consider investing more into research on NHI, implementing nationwide compulsory NHI programmes and establishing a multinational West African collaboration to co-design a sustainable context-specific NHI system based on solidarity, equity and fairness in financial contribution.

## INTRODUCTION

According to the World Health Organization (WHO) in 2022, despite the fact that 75% of national health policies of countries worldwide are directed towards achieving UHC, half of the world’s population still lack access to the health care they need and more than 930 million people worldwide spend at least 10% of their household income on healthcare with approximately 100 million people worldwide impoverished each year due to OOP health expenditure [1]. To attain UHC, countries must strive to achieve better progress on the three main dimensions of UHC which include maximizing the proportions of the population covered, the costs covered and the services included in the benefit packages, all of which could be supported by increasing pooled funds and thus ensuring financial risk protection when accessing healthcare [2, 3].

Financial risk protection has the goal of safeguarding people against catastrophic health expenditure (CHE) and impoverishment due to out-of-pocket (OOP) payment for health services. It forms an integral component of UHC as it ensures that people get access to the essential healthcare services they need without being at risk of financial hardship due to health spending. Furthermore, financial risk protection serves as an interface between a country’s health system and other social determinants of wellbeing [4]. This is evidenced by the fact that, the choice to access health services will be made by an individual if it does not come at the cost of sacrificing other basic needs or imperative social circumstances such as food security, shelter and basic education due to the risk of financial hardship [4, 5]. The devastating effects of OOP health expenditure on financial risk protection have been explored by many economists and researchers in different countries as shown by studies published in Malawi, India, Chile and Spain reporting on CHE and impoverishment [6–9].

The introduction of health insurance schemes in countries thus seeks to avert the financial burden placed on households due to OOP payments for health services [4]. Health insurance is a contract that binds two parties where a policyholder (insured) contributes premiums to a third party payer or a government program (insurer) [10]. Health insurance schemes guarantee that funds collected are pooled to ensure efficient risk sharing among the insured in such a way that the cross-subsidies are from healthy to the sick, rich to the poor, young to the old and those without children towards those with children while eliminating the risk of any single insured member incurring impoverishment as a result of health expenditures [10–13]. In fact, studies conducted in Ethiopia, China and Vietnam have affirmed the protective effect of health insurance schemes against CHE and impoverishment due to health expenditure [14–16].

International studies have reported different systems through which modern public health insurance schemes are operationalized in countries, these include three main schemes: the National Health Insurance (NHI), the Social Health Insurance (SHI) and the Community-based health insurance (CBHI) [17, 18]. The NHI is a compulsory government-run insurance system whereby the government serves as the single third party payer and the policyholder is allowed to individually purchase a health insurance after which funds of the entire population are pooled at a national level [19]. Under the SHI funds for health financing are raised through compulsory premiums deducted directly from the employee’s payroll taxes. Funds are usually pooled and managed by more than one third party payer (mostly public or quasi-public organization) [17, 20]. The CBHI employs the concept of pooling funds for health financing, however funds are normally collected and pooled in a subgroup of the population, mostly at a community level [17, 21].

The health financing mechanisms in West Africa have been known to face many challenges such as general insufficient national income, dedication of majority of general government expenditure to non-health-related sectors, insufficient external revenue for health, high informal sector workforce resulting in limited government tax revenue for health and marked corruption in the public sector [25–27]. The low prioritization of the healthcare sector by governments of the West African subregion continues to undermine the objectives of UHC [25]. The majority of the population end up paying OOP for healthcare services which predisposes them to impoverishment [28].

In the early to mid-2000s, the health insurance system was introduced into West Africa to pool risk in order to address the issue of frequent OOP payments while improving equitable access to healthcare [29, 30]. The NHI scheme is the predominant scheme in most parts of West Africa because other schemes, especially the SHI schemes, are considered less feasible and sustainable in most parts of West Africa because the majority of the population active in the informal sector and have no fixed salaries thus making compulsory deductions from payroll taxes a major challenge [20].

Furthermore, studies report that the poor policy implementation, the administrative burden of premium collection, the inefficient pooling of health insurance funds as well as the inequitable allocation of resources have weakened the health insurance systems and placed an additional burden on the already challenged health financing mechanisms of West African countries [4, 31–33]. In order to improve on the existing NHI systems and facilitate the attainment of UHC in West Africa, it is important to examine the extent to which the current NHI systems contribute to the financial risk protection of insured households in West Africa.

In this systematic review, our research question was; are households in any West African country enrolled under NHI schemes at lower risk of CHE and impoverishment due to health spending as compared to uninsured households? Our main aim was to describe the extent of financial risk protection among households enrolled under NHI schemes in West Africa from published literature and our objectives were; 1. To ascertain the effect of NHI on CHE and impoverishment due health spending in West Africa. 2. To ascertain the proportion of households under NHI schemes incurring CHE and impoverishment due health spending in West Africa. 3. To describe the current NHI schemes and their financing mechanisms in West African countries. 4. To summarize existing recommendations on the development of NHI from published literature and formulate potential strategies to improve the extent of financial risk protection among those currently insured under NHI schemes in West Africa.

## METHODS

### Study design and study registration

Our study was a systematic review conducted and reported following the Preferred Reporting Items for Systematic Reviews and Meta-Analyses (PRISMA) standards. The review protocol was registered with the International Prospective Register of Systematic Reviews (PROSPERO) with ID CRD42022338574 on 28th June 2022.

### Search strategy

After specifying the research question with the PECO (Population, Exposure, Comparison, Outcome) framework (Table 1) and conducting an initial literature review, the final search strategy was developed using keywords, synonyms and Medical Subject Headings (MeSH) terms related to NHI, financial risk protection and UHC in all West African countries in order to increase yield of articles found.

**Table 1:**
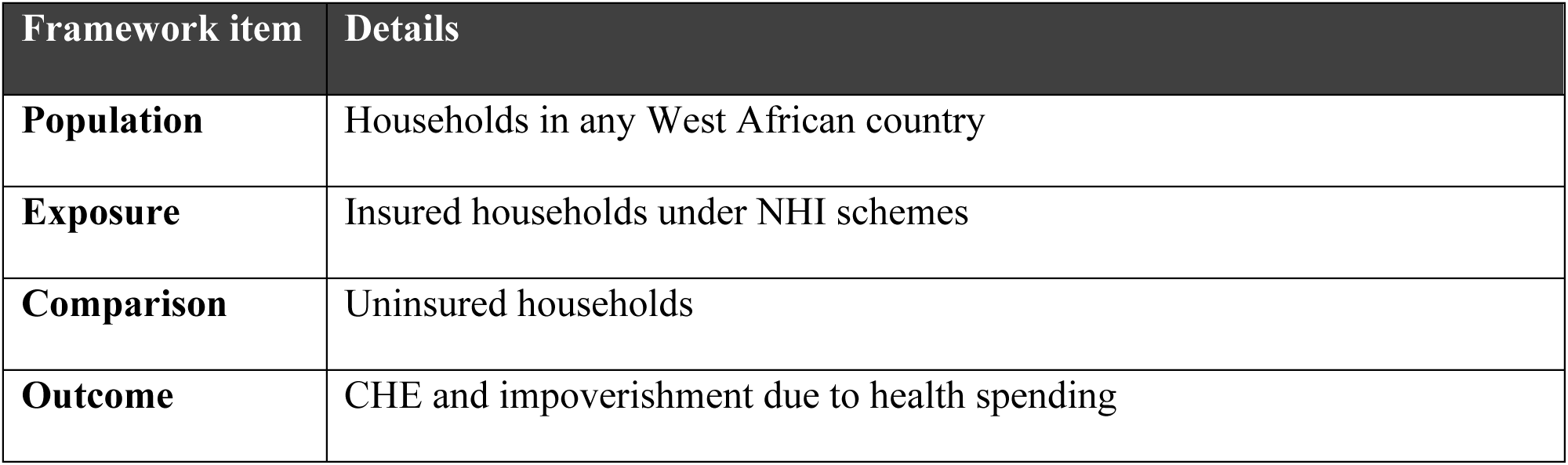
Framework to structure the research question (PECO - Population, Exposure, Comparison, Outcome).

| Framework item | Details |
| --- | --- |
| **Population** | Households in any West African country |
| **Exposure** | Insured households under NHI schemes |
| **Comparison** | Uninsured households |
| **Outcome** | CHE and impoverishment due to health spending |

Four major electronic databases namely PubMed/Medline, Web of Science and Cumulative Index to Nursing and Allied Health Literature (CINAHL) via EBSCOhost and Embase via Ovid were searched. Grey literature was also manually searched for on Google Scholar using advanced search string to identify relevant articles that were not published. The last search of the databases was run on 1st May 2022.

After titles and abstracts screening, citation tracking and snowballing methods were applied on articles eligible for full-text screening to help find more related relevant articles that met the eligibility criteria. The complete search strategies for each electronic database are presented in Supporting Information (SI) 1.

### Eligibility criteria and study selection process

Titles and abstracts of studies were screened for relevance. Full texts of relevant articles were assessed for inclusion applying various eligibility criteria. Studies were considered eligible for inclusion if studies with available full text were:

- observational studies (cohort studies, cross-sectional studies and case-control studies) conducted in any West African country.
- written in English and published between 1st January 2005 and 1st May 2022. This year limitation was applied because the NHI was introduced into West Africa in the early to mid-2000s and as such published articles from 2005 to date yielded more relevant studies to answer the research question [29, 30].
- describing the NHI system in any West African country in relation to revenue raising, pooling of funds and purchasing of services.
- mentioning financial risk protection and/or UHC among populations enrolled into NHI schemes in any West African country.
- assessing CHE and/or impoverishment due to health spending among populations enrolled into NHI schemes in any West African country.
- estimating CHE using either total household expenditure/income or non-subsistence expenditure/income.

Studies were excluded from the review if they:

- were reviews, protocol papers, letters, editorials, conference abstracts and poster presentations.
- had no clear aims and objectives or did not report on ethical approval.
- had households with private health insurance as a comparison group.

For the study selection, the citation of studies identified through the electronic databases including abstracts and full texts were downloaded and imported into Covidence online tool. The Covidence tool automatically removed all duplicates which were confirmed by two independent reviewers (SO and FK). The titles and abstracts of the remaining articles were screened for relevance and for all relevant studies, both reviewers independently assessed the full-text by carefully applying the eligibility criteria. Disagreements encountered in the process of title, abstract and full-text screening were discussed between the two reviewers and resolved by consensus before moving to the next phase. The study selection process was finally presented in a PRISMA flowchart (Figure 1).

**Figure 1:**
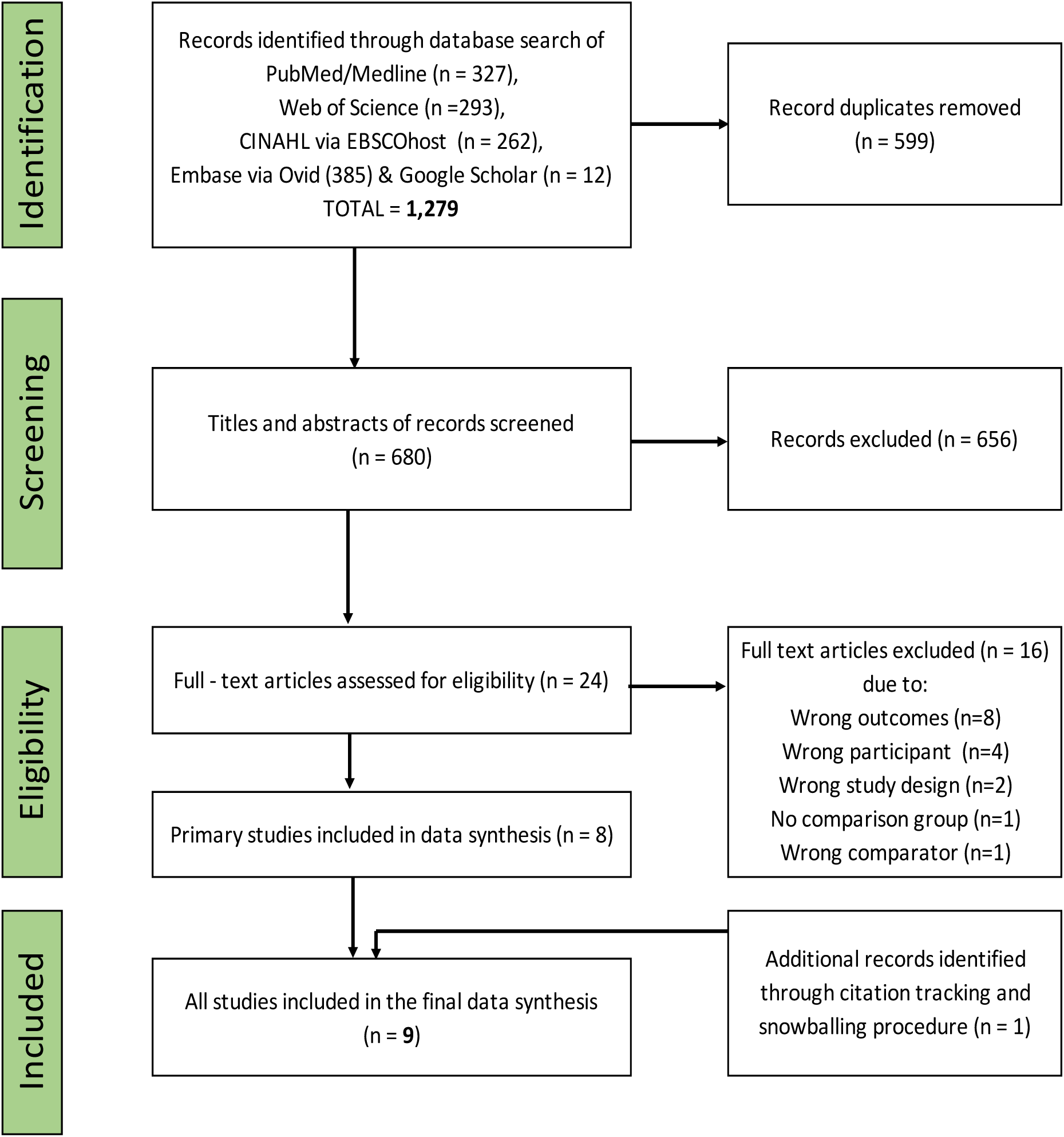
PRISMA flowchart showing the study selection process. Source: Adapted from template provided by PRISMA: http://prisma-statement.org/

### Data Extraction

Data was extracted using a purposefully designed data extraction form. The initial data extraction form was pilot tested using two randomly selected articles of the final included articles and subsequently revised to ensure that all relevant information was captured by the final data extraction form.

Information extracted using the final data extraction form included the publication details (authors, year, title and citation), the study and participant characteristics and the exposure characteristics. The primary and secondary outcome measures, the effect measures as well as additional information from authors were extracted. The exposure of interest was insurance status as defined by household enrolment into NHI schemes or not. Criteria for classifying household as insured was also recorded. The total number of insured households in the sample population, the health insurance type and information on its financing mechanism were collected. Information about confounding variables identified by the authors were also collected.

For the outcome parameters, the mean household income/expenditure and the measure of financial risk protection used in the selected articles were recorded. The respective thresholds used for the measure of CHE and poverty were also collected. The various effect measures used for assessing the association between the exposure and primary outcome were captured. The proportion of CHE and impoverishment at the various thresholds as reported in the selected articles was also recorded. Any information on determinants of CHE and impoverishment as well as health outcomes and service utilization among insured populations were considered as secondary outcomes and recorded so long as they were reported in the articles. Finally, additional information such as limitations, recommendations, study funding sources, concerns about bias and possible conflicts of interest were extracted.

Data was extracted for each study by two reviewers (SO and MAD) independently using the final data extraction form. The primary reviewer (SO) then examined all the extracted data for consistency and accuracy. Discrepancies in extracted data were discussed and resolved by consensus between the two reviewers. The complete data extracted onto the data extraction form is presented in SI 2.

### Quality appraisal

The final included studies were cross-sectional and cohort studies. To assess their quality, the corresponding checklists of the Joanna Briggs Institute (JBI) were selected from the Critical Appraisal Checklists [34–36]. The assessment was carried out independently by two reviewers (SO and FK). Discrepancies between the two reviewers were clarified by discussion until a consensus was reached. The Idostatistics online statistical tool was used to compute the inter-rater reliability (Cohen’s kappa score) and the percentage of agreement between the two reviewers [37]. We set a scoring system with three categories (high, moderate and low) to allow for comparison of the quality of studies in a transparent manner. For cross-sectional studies, a score of four or less was considered low quality, a score of five and six considered moderate quality and a score of seven and eight considered high quality. For cohort studies, a score of six or less was considered low quality, a score of seven to nine considered moderate quality and a score of ten or more considered high quality.

### Data synthesis and analysis

A narrative synthesis was performed and included an iterative process that consisted of two methods: the thematic synthesis for the qualitative data and the Synthesis Without Meta-analysis (SWiM) which provided a more transparent approach for the synthesis of quantitative data [38]. The thematic synthesis was done by assigning codes generated from identifying recurring themes in the data. The NVivo Release 1.6.2 software was used to organize, sort, arrange and summarize qualitative data. The SWiM method on the other hand used vote counting to capture the direction of effect of NHI on CHE and impoverishment reported by the included studies. For each study, for the association between the exposure and the outcome, three possible effects were considered which included, a positive effect represented by “+”, a negative effect represented by “−” and no statistically significant effect (p-value >0.05) represented by “0”.There was heterogeneity in the measurement of exposure (different criteria for classifying households as insured), measurement of outcomes (varying thresholds for CHE and impoverishment) as well as in the measurement of effect measures among the final included studies. These methodological and statistical heterogeneity did not allow for a formal meta-analysis. The collated elements of the narrative synthesis are presented in tables and texts under topics relevant to the objectives of the study (see Results).

## RESULTS

### Study selection

A total of 1,279 studies were retrieved after the electronic database search, however after removing duplicates and screening the titles, abstracts and full texts, 1,270 studies were excluded and nine were included as the final eligible studies [39–47]. Two out of the 1,270 excluded studies were initially added to the final included studies but during the data extraction it was discovered that they did not meet the criteria for the comparison group. The first study used privately insured households as the comparison group instead of uninsured households [48]. In the second study, the entire sample consisted of households enrolled under the NHI and therefore, there were no uninsured households for comparison [49]. Consequently, both studies were excluded. The complete selection process thus yielded nine studies as shown in the PRISMA flowchart in Figure 1.

### Study characteristics

The nine eligible studies were published between 2011 and 2021 of which eight were conducted in Ghana and one in Nigeria. Also, eight studies were cross-sectional studies while one was a retrospective cohort study. From the included studies, five studies collected primary data by administering questionnaires to individuals or households while four studies analysed secondary data (Table 2). The sample size of the households ranged from 2,418 to 16,772 while that of the individuals ranged 196 to 11,617. Five of the studies had their study sample drawn from the general population irrespective of their health status while four studies drew their study sample from surgical patients on ward admission, tuberculosis patients, seriously injured children and employees in federal or state sector jobs (Table 2).

**Table 2:** Summary characteristics of included studies.

| Publication details | Study Characteristics |  |  |  |  |  |  |  |
| --- | --- | --- | --- | --- | --- | --- | --- | --- |
|  | Country/Region | Study objectives | Study design | Study period | Recall period | Sampling unit | Sample size | General health status |
| Flannery, 2020 [39] | Nigeria/Akwa Ibom state | To examine the relationship between enrolment in a NHI scheme and household health expenditures of federal and state workers in Akwa Ibom, Nigeria. | Cross-sectional survey | 2018 | 4 weeks | Individual | 538 | Federal or state sector employees irrespective of health status. |
| Aryeetey, 2016 [40] | Ghana/ Central and Eastern region | To analyze the effect of health insurance on household OOP expenditure, CHE and poverty. | Cross-sectional survey | 2009-2011 | 1 month | Household | 3,128 *<br>3,128 ¶ | General population irrespective of health status |
| Fiestas Navarrete, 2019 [41] | Ghana (data from GLSS 6 ) | To evaluate the effect of health insurance on financial risk protection and utilization among high-risk and vulnerable beneficiaries with and without limited geographic accessibility to care | Cross-sectional survey | 2012-2013 | N/R | Household | 16,744 | General population irrespective of health status |
| Kusi, 2015 [42] | Ghana, Kwaebibirem/ Asutifi and Savelugu–Nanton | To examine the effect of the NHI scheme on OOP health expenditure and how it protects households against CHE. | Cross-sectional survey | 02.2011 – 04.2011 | 4 weeks | Household | 2,418 | General population irrespective of health status |
| Nguyen, 2011 [43] | Ghana/ data from 2007 survey in Nkoranza and Offinso) | To evaluate the impact of the country's NHI scheme on households' OOP spending and CHE. | Cross-sectional survey | 2010 | 12 months | Individuals | 11,617 | General population irrespective of health status |
| Okoroh, 2020 [44] | Ghana/Korle Bu Teaching Hospital (KBTH) | To test the hypothesis that insurance makes a difference in OOP expenditures for surgical care and CHE by comparing rates of financial catastrophe for insured versus uninsured surgical patients at a single institution, the KBTH. | Cross-sectional survey | 01.02.2017– 01.10.2017 | 3 months | Individual | 196 | Patients admitted on the general surgery ward |
|  | Country/Region | Study objectives | Study design | Study period | Recall period | Sampling unit | Sample size | General health status |
| Pedrazzoli, 2021 [45] | Ghana (data of TB patients at public health facilities across Ghana) | To answer the following questions:<br>1) What are the key drivers of costs and catastrophic costs for TB-affected households?<br>2) If NHI scheme is a driver, what would be the expected changes in experienced costs of expanding the national health insurance scheme to all TB patients? | Cross-sectional survey | 2016 | N/R | Individual | 690 | TB patients |
| Stewart, 2021 [46] | Ghana/ Komfo Anokye Teaching Hospital (KATH) | To determine the effect of the NHI scheme on timeliness of care, outcomes, and household CHE of seriously injured children at a tertiary hospital in Ghana. | Retrospective cohort study | 01.01.2015–31.12.2016 | N/R | Individual | 263 | Seriously injured children |
| Kwakyie, 2018 [47] | Ghana (data from GLSS 6) | The general objective of the study was to determine the effect of NHI scheme coverage on CHE among households in Ghana. | Cross-sectional survey | 2012-2013 | N/R | Household | 16,772 | General population irrespective of health status |

**Note:** \* Data collected in the year 2009, ¶ Data collected in the year 2011, N/R-Not Reported
**NHI:** National Health Insurance. **CHE:** Catastrophic health expenditure. **OOP:** Out-of-pocket. **TB:** Tuberculosis.
**GLSS:** Ghana Living Standard Survey

In all nine studies the insurance type was the NHI, however the criteria for classifying a household as insured differed across studies except for two studies where no criteria were reported. Furthermore, aside one study that assessed financial risk protection in terms of both CHE and impoverishment, all remaining eight studies assessed financial risk protection only in terms of CHE. The assessment of CHE was heterogenous across all studies due to the use of different thresholds in the assessment. Characteristics of the included studies are summarized in Table 2 and 3.

### Quality appraisal

The quality of the included studies ranged from medium to high (Table 4). The common issues that affected the quality included no clear statement on the inclusion criteria (n=2), no reliable measurement of the exposure (n=2), no statement on identified confounding factors (n=1), no statement on strategies to address confounding (n=1) and no reliable measurement of outcome (n=1). Also, the included cohort study did not have clear statements on the follow up time, the completeness of follow up as well as reasons for loss to follow up (Table 4). The overall probability of agreement between the two reviewers was judged as moderate (Cohen’s κ = 0.45) while the overall percentage agreement between the reviewers was 88% with the percentage agreement for each question ranging between 75% and 100% (Table 4).

### Effect of NHI on CHE and impoverishment due health spending

The included studies reported different effects of NHI on CHE at different thresholds of household total income (expenditure) and non-food expenditure. In more than half of the included studies (5 out of 9), NHI was reported to have a positive effect (protective factor) on CHE [39–42, 46]. In addition, one of these five studies reported a positive effect of NHI on impoverishment due to health spending [40]. Furthermore, one out of the nine included studies reported both a positive effect and a no statistically significant effect of NHI on CHE at different thresholds of CHE [47]. In two out of the nine included articles, there was no statistically significant evidence that NHI had an effect on CHE [43, 45]. One out of the nine studies did not report on the effect of NHI on CHE [44]. No study reported a negative effect of NHI on CHE. The summary of the effect of NHI and impoverishment due to health spending is presented in Table 3.

**Table 3:**
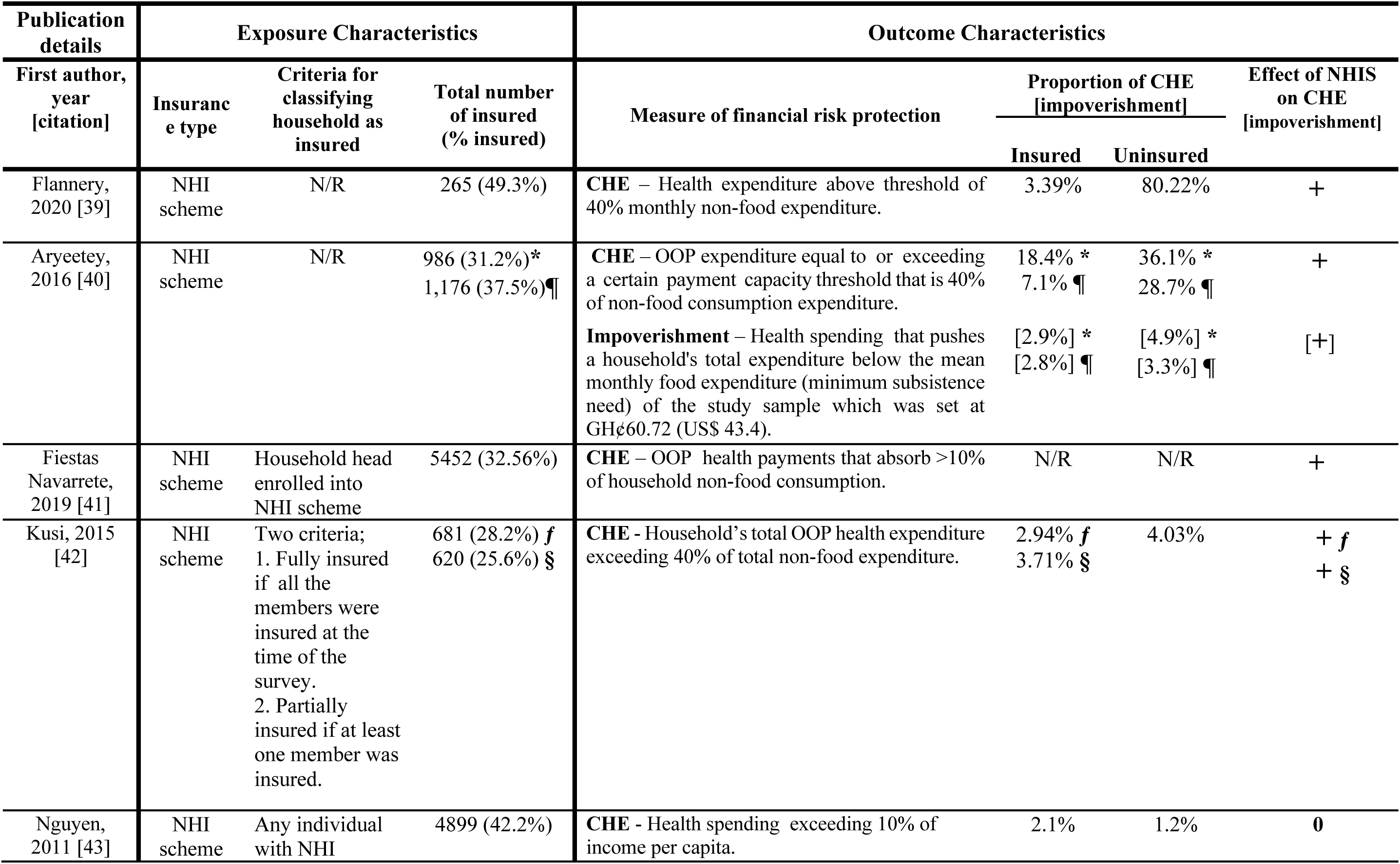
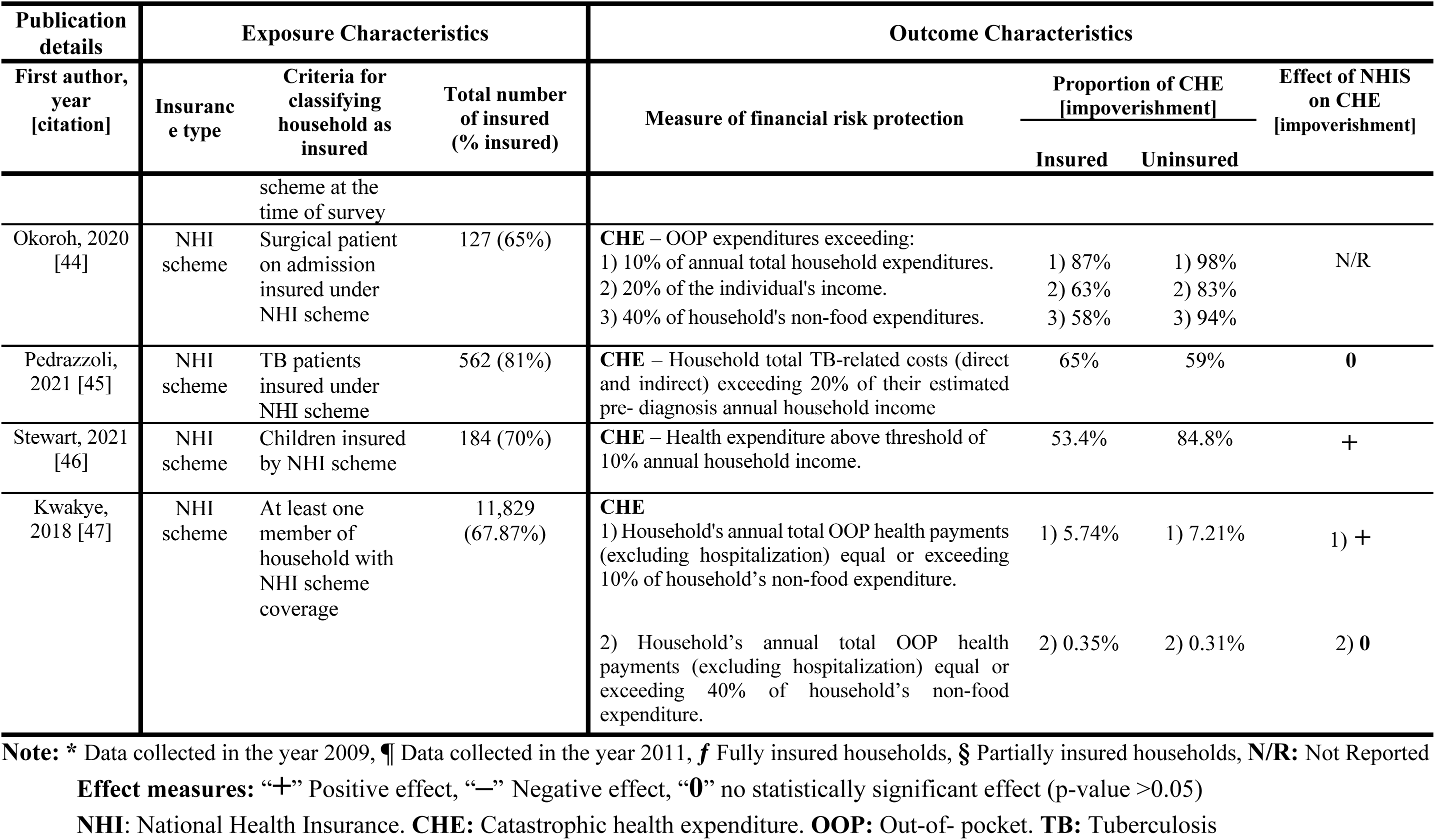
Summary characteristics of included studies

**Table 4:**
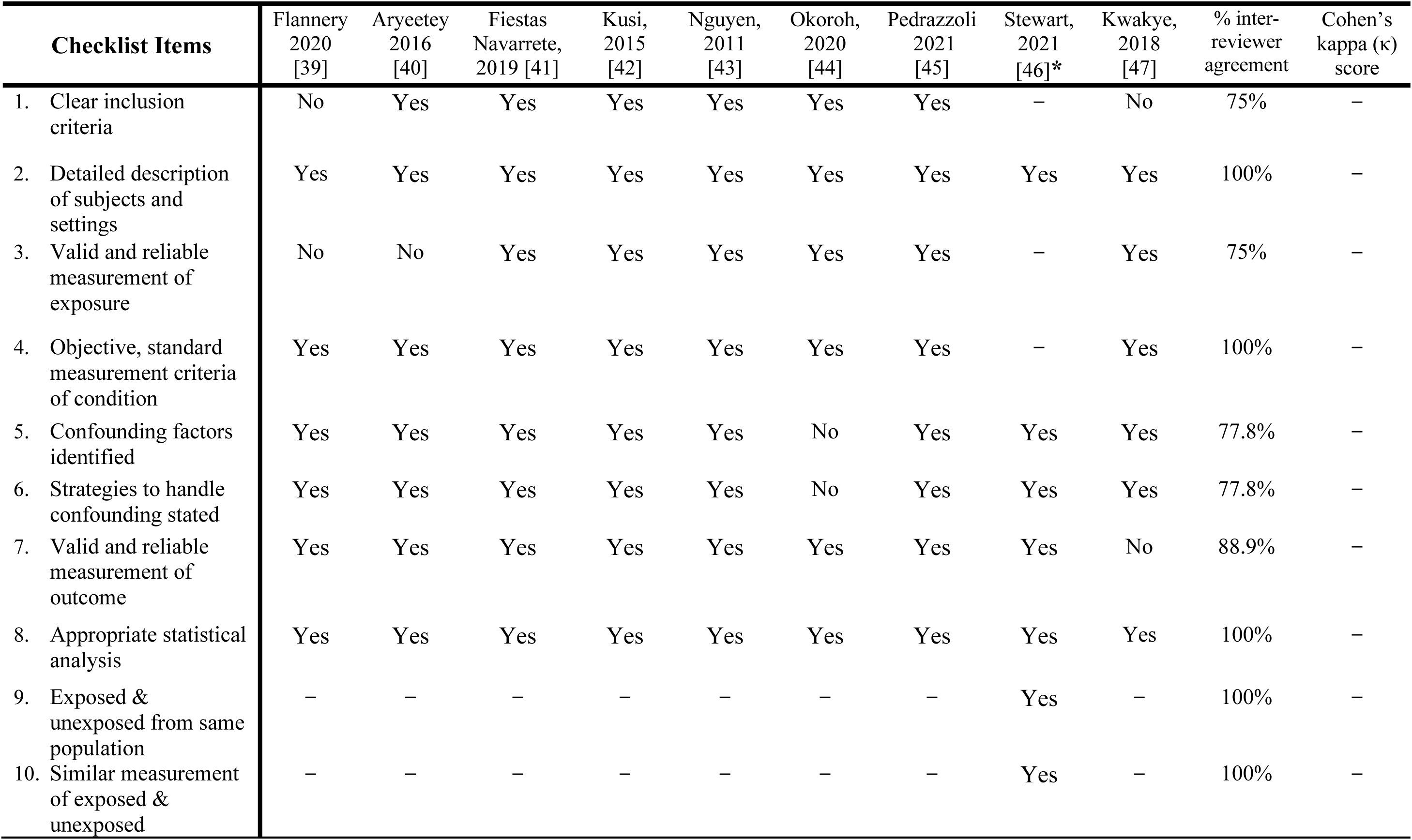
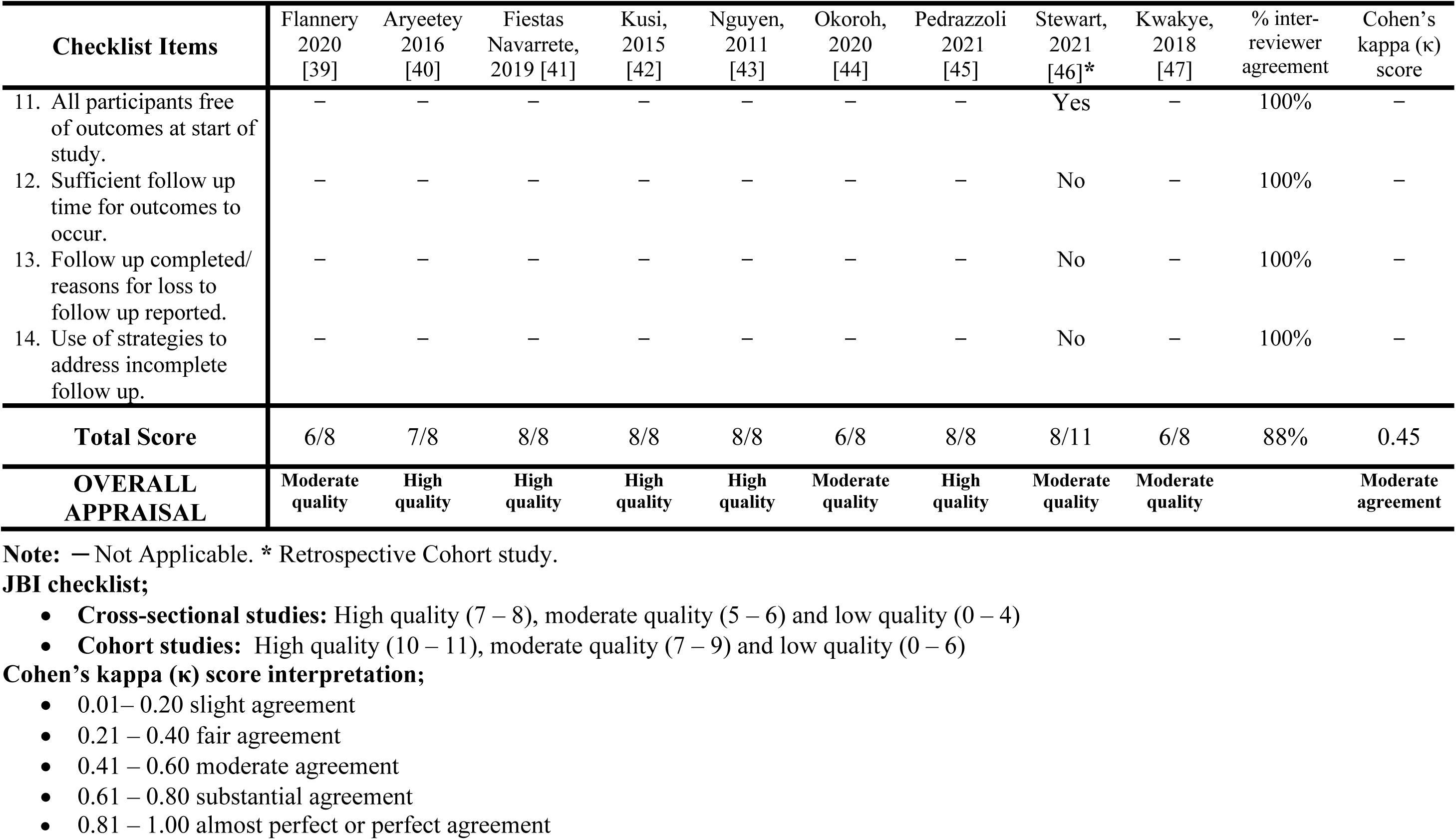
Quality appraisal using JBI checklist

### Proportion of insured households incurring CHE and impoverishment due to health spending

Eight studies assessed the proportion of households that incurred CHE among insured households [39, 40, 42–47]. In seven of these studies, the proportion of households encountering CHE was lower among insured households than uninsured households. In three out of these seven studies, more than 50% of insured households incurred CHE (ranging between 53.4% and 87%) [44–46]. The remaining one out of the eight studies reported a greater proportion of insured households incurring CHE as compared to uninsured households (Table 5) [45]. Only one study reported a proportion of insured households impoverished (below poverty line set at $43.4 per month) due to health spending as 2.9% and 2.8% from data collected in 2009 and 2011 respectively [40]. The summary of the proportion of households that encountered CHE and impoverishment is presented in Table 3.

### Determinants of CHE and impoverishment due to health spending

Six of the included studies reported on determinants of CHE [40, 42, 44–47] while one reported on the determinants of impoverishment due to health spending [40]. The identified determinants were related to the demographic and the health profile as well as the socio- economic circumstances of the households. A large household size, a high number of children under 5 years in a household, a female-headed household and a long distance between a household and a health facility were some demographic determinants of CHE. Furthermore, identified health related determinants of CHE included the health profile of households with at least one member having a chronic NCD, households with any child with severe injuries or any member undergoing surgical procedures or receiving in-patient care. In addition, households with members undergoing diagnostic tests not covered by the Tuberculosis (TB) Programme or NHI schemes such as chest radiography or tests for co-morbidities such as liver function test, undergoing treatment for multidrug resistant tuberculosis (MDR-TB) as well as those with members who make payments for medications were also identified as determinants of CHE. Further, the determinants of CHE related to socio-economic situation of households included patients from poor households, households with no or low level of education and households with unemployed individuals. Finally, the determinants of impoverishment due to health spending reported included large households, households with members who use in-patient services or those with low level of education as well as unemployed members.

### Effect of NHI on health outcomes and health service utilization

One study reported health outcomes as mortality in terms of number of insured and uninsured patients dying while on admission. In this study, three cases of mortality were reported out of the 184 insured patients while no case of mortality was reported out of the 79 uninsured patients. However, the association between NHI and patient mortality was not assessed in the study [46]. Health service utilization among households with insurance was reported by three studies but only one study reported a statistically significant positive effect between NHI and health service utilization and concluded that households with NHI were more likely to utilize health services compared to uninsured [41]. The two remaining studies reported an increased frequency of health service utilization among households with NHI as compared to uninsured households however no statistical analysis was performed to determine the measure of effect between NHI and health service utilization [39, 44].

### Financing mechanism of NHI systems in Ghana

As reported from the eight included studies conducted on Ghana [40–47], the NHI scheme in Ghana established in 2003 relies on diversified set of funding sources which includes a 2.5% levy on goods and services under the Value Added Tax (VAT) which contributes about 70% of the pooled funds. Additionally, 2.5% of the Social Security and National Insurance Trust (SSNIT) contributions are automatically deducted from salaries of formal sector workers and contributes about 20-25% of the pooled funds. The remaining 5% of funds come from voluntary income adjusted premiums by adults (18–69 years) in the informal sector.

However, portions of the Ghanaian population are exempted from payment of premium and they include pregnant women and their newborn, children under 18 years whose parents are enrolled under the NHI scheme, SSNIT pensioners, the elderly over 70 years, impoverished populations and individuals with mental disorders. Those exempted from premiums make up about 60% of the entire insured population enrolled under the NHI scheme and enjoy the full benefits that other premium paying members are entitled to.

Furthermore, based on the description from the included studies, the benefit package under the Ghana NHI scheme covers about 95% of the country’s health conditions and comprises of outpatient care including medications and laboratory diagnostics, in-patient services, surgical and obstetric care, treatment of cervical and breast cancers, basic oral health and ophthalmological services and emergency and trauma care. Despite the extensive conditions included in the benefit package, some services are still excluded under the NHI scheme and these services consist of other cancer treatment aside cervical and breast cancers, dialysis for chronic renal failure, antiretroviral medications and specialized care, medications and diagnostics for the management of trauma such as advanced diagnostic imaging, prosthetics, rehabilitation and mortuary fees.

Moreover, the included study reported that the NHI agency is the regulatory body of the NHI scheme in Ghana and is responsible for setting the premium and registration fees, pooling funds, negotiating benefit packages, accrediting and paying health service providers and ensuring quality service from health service providers. The administrative function of the NHI agency which includes the collation of insurance claims is decentralized to the district level. Finally, the service providers receive payment within four weeks of submission of their insurance claims [40–47].

### Financing mechanism of NHI systems in Nigeria

The study conducted on Nigeria reported that the NHI scheme was launched in 2005 and is made up of three packages with different mechanisms of financing. These packages include the Formal Sector Social Health Insurance Programme (FSSHIP), the Urban Self-Employed Social Health Insurance Programme (USSHIP) and the Rural Community Social Health Insurance Programme (RCSHIP).

This study focused on FSSHIP and as such described its financing mechanism [39]. It reported that the funding for the FSSHIP comes from an annual premium that is 15% of an employee’s annual basic salary which is a shared contribution between the employee (5%) and the employer (10%). Public and private sector workers, the armed forces, the police, para-military organizations and tertiary students are the main targets of the FSSHIP however this NHI scheme is open to voluntary contributors. No groups are exempted from this health insurance scheme.

Furthermore, the study mentioned that the benefit package under the FSSHIP in Nigeria includes preventative care (immunizations), out-patient services, in-patient care up to 15 days per year, obstetric care for up to four live births, eye and dental care, prostheses and prescribed medications and diagnostic tests available on the national recommended lists. Excluded services under the FSSHIP comprise of palliative care for terminal illnesses, antiretroviral medications, chronic diseases (diabetes and hypertension), renal dialysis and spectacle and contact lens provision.

It was also reported that the Health Maintenance Organizations (HMOs) are the main organizations responsible for collecting premiums, pooling funds, paying for health services as well as ensuring quality of care by health service providers under the FSSHIP. The HMOs are usually publicly or privately owned limited liability companies who aim at providing cost-effective healthcare delivery. Payment of providers under the FSSHIP depends on whether care received by insured is at primary healthcare facility, secondary or tertiary healthcare facility: for primary care payment is by capitation whereas for secondary and tertiary care, payment for services provided is based on fee-for-service [39].

### Summary of limitations of included studies

Limitations reported in the final included studies were related to the study design, information recall, outcomes assessment, identification of confounders and generalization of findings to the entire population of countries. The cross-sectional nature of most of the study designs gathered data at one point in time and made it impossible to follow up on households to ascertain if they incurred recurrent CHE. Furthermore, the retrospective cohort study had similar limitations in that, it did not follow children longitudinal even though seriously injured children may require numerous health visits within the course of their treatment which will be associated with OOP health expenditure which predispose to CHE.

Studies also observed that it was difficult for certain households where members were informal sector workers or had unstable or seasonal income to accurately recall their incomes. One study reported a short recall period as a limitation because it may underestimate health services utilization and subsequent OOP payments which may be associated with CHE. Some studies used the national median household expenditure to estimate CHE which may not reflect the median annual household expenditure of the study sample and give imprecise results.

Another limitation mentioned was that studies did not explore if households had to borrow or sell assets to make payment for health services and how they coped with CHE and its impoverishing effects. Possible confounding factors such as type of illness and disabilities among household members, existing health care systems and availability of social protection policies were also not identified in certain studies though these may influence the effect of NHI on CHE. The results of some studies were also restricted to subgroups of the population such as formal sector employees, patients on admissions and certain districts. This limited the generalizability of the results to the entire population of the various countries. Lastly, one study reported a selection bias towards insurance with insurance coverage among study participants two times more than that of the national average as such limiting the extrapolation of the findings to the national level.

### Summary of recommendations by included studies

The included studies recommend numerous strategies to develop NHI schemes to protect households against CHE and impoverishment due to health spending. Most studies advocated the need for future research to better explore factors that allow for an effective and well-functioning NHI scheme. Studies recommended impact evaluations using other study designs to clearly identify the factors of NHI schemes that positively impact OOP payments. One study mentioned the importance of conducting benefit incidence analysis to document how different households with different wealth status benefit from NHI schemes. In-depth case studies were also recommended to explore how NHI schemes are operationalized and to assess the quality of care received by households enrolled under NHI. Advocacy was made for more qualitative studies to explore the reasons for low NHI enrollment rates among rural dwellers to help extend coverage to include these populations.

Additionally, gathering of evidence to identify the exact OOP health expenditure which predisposes households with NHI to CHE was recommended by some studies. In relation to TB management under NHI schemes, it was recommended that further investigations be made into why the current NHI does not effectively protect against CHE among insured TB patients. Other studies recommended development towards a more inclusive and equitable NHI scheme that will ensure that populations that are vulnerable due to their socio-economic and geographic characteristics will also benefit from the financial risk protection as well as the health services utilization effect of NHI schemes.

Some studies recommended that physical access to health facilities should be improved by considering transportation cost as part of the benefit package of NHI schemes. It was also recommended that measures be put in place to ensure strict adherence of health facilities to benefit packages under NHI schemes. Also, a study reported that the rates of payment of health facilities by insurance agencies for surgical care under the NHI scheme should be improved. Finally, to better understand the overall impact of NHI on CHE, it was recommended that studies with reliable nationwide datasets be conducted to get results that can be extrapolated to the general population to guide policymakers in decision making for the effective implementation and operationalization of NHI schemes nationwide.

## DISCUSSION

At the time of this review, this appeared to be first systematic review to ascertain the extent of financial risk protection among households enrolled under NHI schemes in the entire West Africa as well as to summarize potential learnings and recommendations for countries in the subregion. Overall, two-thirds of the included studies revealed evidence suggestive that enrollment into the NHI was a protective factor against CHE however these studies also reported that a percentage of households enrolled under NHI schemes continue to incur CHE and impoverishment due to health spending with one-third of the included studies [44–46] reporting more than 50% of insured households incurring CHE.

### Study characteristics

The small number of included studies suggests a scarcity of published literature exploring the impact of NHI schemes on financial risk protection in West Africa. The included studies were observational studies consisting of either cross-sectional or retrospective cohort studies. This finding is consistent with similar studies carried out in Iran, India and other LMICs [50–52]. In all these studies, the observational study design was favored in place of Randomized Controlled Trials (RCTs) to evaluate the effectiveness of public health policies although RCTs are well established in proving the effectiveness of interventions [53]. Observational studies are more feasible compared to RCTs because in the evaluation of public health policies like NHI schemes it is controversial and would be unethical for a state to randomly assign a population into insured and uninsured groups, when the uninsured group is at a high risk of undesirable outcomes such as CHE and impoverishment due to health spending [54]. Moreover, in public health policy research where findings of studies might be used for nationwide decision making, RCTs are not feasible since findings of RCTs cannot be easily extrapolated to the entire population due to the limited sample used [53]. Indeed, one study in rural Cambodia applied the RCT design to evaluate the effect of health insurance on economic outcomes, health care utilization and health outcomes among publicly insured and privately insured groups. However, this research was solely carried out by a private insurance company with a limited sample size and as such it would not be feasible to generalize the findings of this study at country level [55]. These are possible reasons why the final included studies in this review were observational studies.

Despite the fact that the focus of this review was on the West African subregion, almost all of the final included studies were from Ghana with one from Nigeria. This finding suggests not only an abundance of studies from Ghana but also a geographic gap in the available literature from French speaking countries. Since a rigorous and transparent study selection process was employed for this systematic review, it is unlikely that this finding is due to bias. The paucity in literature from French-speaking countries could be due to the fact that out of the 15 countries in West Africa only 5 countries (Gambia, Ghana, Liberia, Nigeria and Sierra Leone) have English as their official language with the remaining predominantly French-speaking countries. Therefore, per the protocols of this study, only studies published in English were eligible for inclusion with French literature being excluded.

Furthermore, among the English-speaking countries, only Ghana and Nigeria are LMICs with the remaining three LICs therefore enough funds may be available in Ghana and Nigeria for investment into health systems research compared to the other countries [24]. Moreover, a study conducted in 2020 which summarized the availability of evidence for the implementation of health interventions in West African countries showed an abundance of publications from Ghana and Nigeria [56]. These factors could have accounted for the high yield of relevant literature from Ghana and to a lesser extent Nigeria as compared to other countries in West Africa.

Additionally, although for this review the inclusion criteria were studies published between 2005 and 2022, the final included studies were published between 2011 and 2021. This finding might have been due to the fact that between 2005 and 2011, there were no studies that evaluated the association between NHI schemes and CHE and impoverishment due to health spending in West African countries. Furthermore, the absence of relevant published literature in 2022 may have been due to the dedication of resources in research toward publications relevant to the coronavirus disease during the pandemic.

### Effect of NHI on CHE and impoverishment due health spending

The financial risk protection effect of NHI was evidenced in this review when majority of the included studies (6 out of 9) reported a protective effect of NHI on CHE with one study of these additionally reporting a protective effect of NHI on impoverishment due to health spending [39-42, 46, 47]. These results were consistent with a systematic review on the effect of NHI on financial risk protection conducted in 2019 among LMICs which reported a protective effect of NHI on CHE in majority (9 out of 14) of the included studies [51]. The consistency of this protective effect of NHI on CHE suggests that households enrolled into NHI schemes are more likely to have reduced or no OOP health expenditure compared to uninsured households therefore NHI households are better protected against CHE and impoverishment due to health spending.

These findings regarding a protective effect of NHI on CHE were observed at different thresholds of CHE which included threshold exceeding: 40% of a household’s non-food expenditure [39, 40, 42], 10% of a household’s non-food expenditure [41, 47] and 10% of a household’s annual income [46]. However, other included studies found non-significant effects of NHI on CHE at thresholds exceeding 10% of income per capita [43], 20% of annual household income [45] and 40% of a household’s non-food expenditure [47]. A systematic review conducted among LMICs also reported protective and non-significant effects at different thresholds of CHE [51]. Overall, there are inconsistencies regarding the thresholds of CHE at which a positive effect of NHI can make a significance difference among insured households in LMICs. This may be because depending on the socio-economic status of a household, small amounts of health expenditure will be catastrophic for the household if the set threshold of CHE is low. For instance, a poor household will have an increased risk encountering CHE after making OOP payments if a threshold of 10% is used while this same household will not encounter CHE at a threshold of 40% [57, 58].

### Proportion of insured households incurring CHE and impoverishment due health spending

Despite the protective effect of NHI on CHE, most of the included studies (8 out of 9) reported a proportion of households insured under NHI schemes experienced CHE at various thresholds [39, 40, 42–47]. This was also found by a household survey conducted in Iran in 2015 which reported a proportion of 5% of insured households incurring CHE at the 40% threshold [59]. A reason for the consistency in findings could be because just like Ghana and Nigeria, Iran is a LMIC and insured households may be subjected to high OOP payments which will predispose to CHE [24, 50]. Furthermore, in LMICs cost of indirect payments such as cost of transportation to health facility (especially in cases of emergency) as well as feeding while on admission may not be accounted for in benefit package of NHI schemes may result in CHE among insured households [51].

In three of the included studies, the focus was on patients on admission on the general surgery ward, TB patients and seriously injured children with 87%, 65% and 53.4% of the insured households incurring CHE respectively [44–46]. These findings were observed at the 10% and 20% thresholds of CHE and were higher compared to the recent WHO Global Monitoring Report on Financial Protection in Health that reported that in 2017, in the African region, 8.4% and 2.0% of households incurred CHE at the 10% and 25% thresholds respectively [60]. This supports the idea that insured households with members undergoing general surgery and TB treatment as well as those with seriously injured children are likely to make OOP payments leading to CHE even though these conditions are included in the benefit package of the current NHI schemes in parts of West Africa [39–47]. Overall, the increased proportion of insured households incurring CHE in parts of West Africa suggests a possible weakness of the NHI in its current state in guaranteeing financial risk protection of the insured household and this may be a significant barrier to achieving UHC in West Africa.

Furthermore, one of the included studies reported that 2.9% and 2.8% of insured households made health expenditure that pushed them below the poverty line set at $43.4 per month in 2009 and 2011 respectively [40]. On the other hand, the WHO Global Monitoring Report on Financial Protection in Health stated that in 2017, the proportion of households in the African region falling below the internationally standardized poverty line of $3.20 per day due to health expending was 1.1% [60]. However due to the inconsistencies in the set poverty lines and possible externalities such as inflation, comparisons on the proportions reported by the included study and that of the WHO report could not be made. Hence, to facilitate a better comparison of data on impoverishment due to health spending, researchers should endeavor to employ standardized context-specific poverty lines while conducting research in this field.

### Determinants of CHE and impoverishment due to health spending

This systematic review reported that households with members who have low level of education or are unemployed as well as households already living below the poverty line are more likely to incur CHE and impoverishment due to health spending. These findings were consistent with evidence from cross-sectional surveys conducted in Zambia and Myanmar which reported that these same determinants played a key role in CHE and impoverishment due to health spending [61, 62]. The consistency in findings supports the idea that in LMICs where the illiteracy, poverty and unemployment rates are high [63–65], households tend to incur CHE and impoverishment due to health spending. A possible reason is that poor households and households with unemployed members are likely to encounter CHE even at low OOP payments due to their low or no income whereas households with low education levels may not even have the required knowledge to make informed decisions such as purchasing health insurance premiums to protect themselves against CHE. Moreover, these three determinants of CHE and impoverishment due to health spending are interlinked in that, with low level of education, household members may be unable to gain employment which will eventually lead to poverty. Additionally, already impoverished households will be unable to provide their members with the necessary education which will help them secure jobs leading to a vicious cycle of illiteracy, unemployment and poverty. Overall, this suggests that general social protection measures such as poverty alleviation, employment as well as educational schemes will play a vital role in guaranteeing financial protection in the drive towards UHC.

Furthermore, this review also showed that households with members with chronic NCDs or on hospital admission were more likely to incur CHE. This finding was in line with studies conducted in India and Tanzania which reported households with these same health profile to be at increased risk of incurring CHE [66, 67]. The finding supports the fact that although the burden of NCD is on the rise in LMICs, there is a general inadequacy in the health insurance coverage for chronic NCDs as reported in this review and the supporting evidence from India and Tanzania and this places financial burden even on insured households [66, 67]. Moreover, the prolonged contact with healthcare providers during management of chronic NCDs and patients on hospital admission as well as the high cost of medications worsens the financial burden on households. Indeed, patients with chronic NCDs are at risk of developing complications like stroke and end-stage renal diseases that will require long-term in-hospital care with subsequent OOP payments resulting in CHE [68].

This study identified multiple determinants of CHE related to the household characteristics such as large household size, high number of children under 5 years in households, long distance to health facility and female-headed household. Firstly, this review observed that increasing household size was associated with an increased likelihood of incurring CHE. This was consistent with a study conducted in Kenya in 2016 which reported that household size exceeding the average size of 5 members were more likely to incur CHE [69]. Kenya, like the West African countries covered in this review are all tropical countries and at higher risk of infectious diseases like Malaria which can affect multiple household members within the same time period [70]. Therefore, the larger the household, the higher the likelihood that multiple household members will contract an infectious disease and require healthcare within the same time period and this may result in high OOP payments with subsequent CHE at household level.

This review also reported that households with more children under 5 years were at risk of incurring CHE. A cross-sectional study carried out in Iran also reported that households with children under 5 had a greater risk of encountering financial burden due to health spending [71]. Both studies support the idea that children under 5 are prone to childhood illnesses and injuries and as such will require more healthcare services. Therefore, the more children under 5 in a household, the greater the likelihood of making OOP payments which may lead to CHE.

Additionally, it was observed from this present review that a longer distance between household and health facility was associated with a higher risk of incurring CHE. This same finding was reported in a study conducted in India in 2017 [72]. The consistency in findings suggests that in parts of LMICs, households have to travel long distances to access health facilities and this journey is associated with high transportation costs which may predispose households to incurring CHE. More so, this physical barrier in accessing health facilities also undermines the drive towards UHC.

Lastly, this review reported that female-headed household were at risk of incurring CHE. This finding was consistent with a cross-sectional study conducted in Ethiopia which also reported that female-headed households were indeed at greater risk of encountering CHE [73]. The consistency in the findings supports the fact that more efforts still need to be put into female empowerment in developing countries [74]. This is because poor socio-economic status among female-headed households and the general lack of female empowerment to make decisions such as purchasing NHI predispose these households to the devastating financial burden of OOP payments and CHE.

Overall, the determinants of CHE and impoverishment due to health spending are significant barriers to UHC and should be taken into consideration during the implementation and operationalization of NHI schemes in West Africa in order to guarantee financial risk protection for households.

### Effect of NHI on health outcomes and health service utilization

One study observed mortality related to NHI status and reported a higher mortality amongst hospitalized insured patients (3 cases) compared to uninsured patients (no cases) over the same period of time for patients undergoing treatment for severe injuries [46]. The opposite relationship was reported by a retrospective review conducted in the United States of America amongst severely injured patients where those insured were found to be significantly better protected against mortality than those uninsured [75]. However, the study included in this systematic review did not perform statistical analysis to ascertain the association between NHI status and mortality and therefore, the reported findings in this study could be partially explained by severity of the injuries or possible complications. In addition, the apparent contradictory findings between the two studies might be due to differences in the health financing system, the healthcare system, the quality of care of patients as well as other factors outside the healthcare system.

In three of the included studies [39, 41, 44], it was observed that insured household utilized health services more frequently than uninsured households, however, only one study among these three reported statistically significant evidence to this observation [41]. This finding was in line with a cross-sectional study conducted in Vietnam in 2020 which found that enrollment into health insurance increased the likelihood of households utilizing outpatient services [76]. The consistency in findings supports the idea that since insured households make no or reduced OOP payments at health facilities, they readily seek healthcare compared to uninsured households who bear the entire cost of healthcare themselves. Although this increased health seeking behavior among insured households is desirable in the drive towards UHC, there is a potential risk of moral hazard due to overutilization of health services. Thus, improvements of the existing NHI schemes should also consider measures to address moral hazard to ensure sustainability of the schemes.

### Financing mechanism of NHI systems in Ghana and Nigeria

From this review, it was observed that in the Ghanaian NHI system, formal sector workers make compulsory contributions while informal sector workers were allowed to voluntarily purchase insurance premiums. Furthermore, in Nigeria, different NHI schemes were established for different subgroups of the population who share common characteristics. Here, pooling of funds takes place at subgroup level for instance among federal and state workers (FSSHIP). These West African NHI systems differ from the German SHI system where payment of premiums is compulsory for a large part (90%) of the population and funds are equitably pooled based on the concept of solidarity where the rich and healthy subsidize for the poor and sick [77, 78]. Despite the fact that the German SHI system is from a developed country, it can be used to make comparisons because as one of the oldest and well-established health insurance systems, it has formed the basis for many health insurance systems across the world [12]. The current mechanisms of raising funds for health services in some West African countries lead to inequity and inequality in pooling of funds. This is so mainly due to the voluntary and subgroup nature of purchasing NHI premiums resulting in ineffective pooling and subsequent low availability of funds for payment of health services.

Additionally, as evidenced in included studies, the Ghanaian NHI system exempted up to 60% of the insured population from payment of premiums. The exempted group consisting of pregnant women, children under 18 years, elderly over 70 years and impoverished populations are considered vulnerable populations and are excluded from payment of premiums irrespective of their capacity to contribute. In comparison with the Nigerian FSSHIP that did not have any groups exempted from payment of premiums just like the German SHI [77], the high number of exemptions in Ghana places a burden on those who pay premiums and this results in inequitable pooling of funds with consequently low funds available for financing health services.

All included studies mentioned a wide range of services included in the benefit package of the NHI schemes which was highly desirable since it is in line with the drive towards achieving UHC. However, both the Nigerian and Ghanaian NHI schemes reported that numerous conditions such as diabetes, hypertension, certain cancers and chronic renal failure are not included in the benefit packages. These conditions excluded are mostly chronic NCDs and considering that current evidence affirms an increasing burden of such conditions across the African continent [79, 80], this finding that the Ghanaian and Nigerian NHI schemes are excluding these chronic NCDs from their benefit package greatly undermines the financial risk protection effect of NHI and can be a major hinderance to the drive towards UHC.

The NHI agency which regulates the NHI scheme in Ghana decentralizes its administrative function to the district level. Evidence from Pakistan suggests that decentralization of the healthcare sector to subnational levels encourages the growth of government measures to improve health service delivery [81]. Therefore, it can also be considered that in Ghana, the decentralization of the administrative functions can contribute to improving the implementation and operationalization of NHI schemes by facilitating the collation and vetting of insurance claims from health facilities, promoting accountability of subnational health services as well as encouraging enrollment into NHI schemes. However, a major disadvantage of decentralization is that high costs of human resources and infrastructure that arise from having various district offices across the country places a financial burden on the public funds of the country. Thus, to ensure the sustainability of NHI schemes, it is empirical for governments to consider innovative ways to fund a decentralized administrative system. In addition, employing digital technology systems in a decentralized organization will play a significant role in simplifying the administrative procedures.

The reimbursement of health facilities for services provided is a vital component of NHI schemes but the included studies from Ghana did not give much information on this. The study from Nigeria reported that the mode of payment for health services depends on which health facility level patients seek care from. This consists of capitation payment at primary care level and fee-for-service at the secondary and tertiary levels. These payment methods differ from that of Germany where payment for ambulatory services is based on fee-for-service [82] and that of hospital services is based on aG-DRG (Diagnostic-related groups) where diagnoses requiring similar treatments are classified into groups and the number of healthcare workers required to manage these conditions are also accounted for before reimbursements are made to the hospitals [83]. The inconsistencies in the method of reimbursement of health facilities between both countries could be due to the existing health policies and health systems organization as well as payment negotiations among other factors. Moreover, the capitation has been found to increase the tendency for healthcare facilities to provide poor quality care while fee-for-service has been linked to escalation of costs [84]. Evidence suggests that the aG-DRG mode of reimbursement for hospitals which as of 2022 is the new DRG system for Germany plays a key role in promoting quality care and ensuring equity in the payment for health services [83, 85, 86]. This new aG-DRG system came into the forefront of health financing discussions in 2021 during the Corona Pandemic which exposed the weakness in the previous DRG system that did not take into account the workforce required to manage patients [87]. Hence, for the improvement of reimbursement of health facilities under NHI schemes in Nigeria and West Africa as a whole, not only should past experiences or systems be considered but more importantly, current trends as well as cultural preferences, perspectives and views of health professionals should be taken into account so as to develop a robust and sustainable NHI system.

### Recommendations

Based on the findings of this systematic review, the following recommendations on future research focus and policy strategies to improve NHI schemes will be vital in ensuring financial risk protection to facilitate the drive towards UHC in West Africa. Since majority of West African countries are French speaking countries, future systematic reviews should include French as an inclusion criterion to capture relevant articles published in French. This will allow for a more comprehensive exploration of the financial risk protection effect of NHI schemes in West Africa. Furthermore, nationwide surveys should be carried out to explore the extent to which existing NHI schemes protect against CHE and impoverishment due to health spending, this will allow for easy generalizability of the results and enable policymakers to make decisions and implement interventions that include the entire population. It is also recommended that researchers have a common criterion for describing household as insured as well as having a common context-relevant threshold for measuring CHE. These common measures will allow for easy comparison and analysis of results to draw relevant conclusions. Future research on financial risk should also consider measuring impoverishment due to health spending in combination with CHE because governments and policymakers are likely to be more sensitive to poverty measures and react accordingly. Continuous studies should also be conducted on the benefit package of NHI schemes with respect to the prevailing disease conditions associated with CHE and impoverishment due to health spending in countries. This may help introduce benefit packages that are not only context-relevant but also effectively protect against CHE and impoverishment due to health spending.

In relation to the policy recommendations, premium contributions should be compulsory for the entire population of countries for NHI schemes to effectively pool sufficient funds for health services. Furthermore, exemptions of payment of premiums should be limited to impoverished populations who would be identified through a robust registration process for these vulnerable groups. In the drive towards financial risk protection and UHC, social protection policies should be rolled out to improve the socio-economic circumstances of the citizens of countries. Also, administrative processes such as vetting of claims should be digitized and decentralized to facilitate effective collation and efficient analysis. Additionally, to reduce the risk of moral hazard, strategies such as well-designed payment of deductibles and coinsurance are recommended. Capacity building and sufficient resource allocation to the field of research focused on financial risk protection and UHC is recommended to equip policymakers with the evidence needed for decision making. Finally, West Africa is under one economic community (ECOWAS) and has a regional health organization known as the West African Health Organization (WAHO) that is responsible for ensuring and promoting the health of citizens and fostering regional governance in health through the investment in health systems research in West Africa [88]. Due its functions, WAHO is in a desirable position to work together with ECOWAS member states to co-design a robust context-specific NHI scheme that could be adopted and implemented by member states. These recommendations complement the aforementioned recommendations summarized from the included studies (see Results) and will overall contribute to improving the financial risk protection effect of NHI schemes and facilitate the drive towards UHC.

### Limitations of the study

Despite the robust methodology of this review, several limitations have to be mentioned. First of all, the findings are based on only nine studies with majority of the studies from Ghana and as such these findings cannot be generalized to all countries in West Africa. Furthermore, only two of the nine included studies had nationally representative data which also limits the generalization of the overall findings. Moreover, because of the timeline set for this study, articles published after 1st May 2022 were not included and as such results presented in this systematic review may not reflect recent developments after this date. Additionally, since only one study reported on impoverishment due to health spending, conclusions drawn on this measure of financial risk protection cannot be generalized to the entire West Africa. Another limitation was the high degree of heterogeneity among included studies due to difference in the criteria for classifying households as insured as well as the thresholds for accessing CHE. Finally, the nine included studies encountered several limitations which may be relevant for future studies and are summarized under the Results section.

## CONCLUSION

This systematic review provides evidence that households insured under NHI schemes in some West African countries (Ghana and Nigeria) are more likely to be protected against CHE and impoverishment due to health spending compared to uninsured households as reported in six out of the nine included studies. However, households insured under NHI schemes continue to incur CHE and impoverishment due to health expenditure with three out nine studies reporting more than 50% of insured households incurring CHE. Reasons for this are numerous and interlinked and can be related to households’ demographic and socio-economic characteristics as well as their health profiles. The most relevant determinants of CHE and impoverishment due to health spending identified among households were poverty and unemployment, presence of chronic diseases and indirect OOP health expenditure such as transportation costs to health facilities. To protect households in West African countries against the devastating effects of financial loss due to health spending, further research using nationally representative data needs to be conducted to ascertain the extent of NHI schemes on financial risk protection in West African countries in order to have relevant data to guide evidence-based decision making. Governments should also consider focusing on rolling out compulsory nationwide NHI premium contributions while implementing effective social protection schemes to reduce the incidence of CHE and impoverishment due to health spending. UHC in West Africa can be made attainable and the role of a multinational collaboration between West African countries to co-design a sustainable context-specific NHI system based on solidarity and equity will go a long way in advancing the drive towards UHC.

## Supporting information

Search strategies: This is a description of our comprehensive list of search strings used to identify all studies in the various electronic databases

Complete data extraction form: This contains the complete data extracted from the final included articles in this review.

PRISMA Checklist

## Data Availability

All data has been provided as part of the submitted manuscript

## AUTHOR CONTRIBUTIONS

Conceptualization: Sydney Odonkor, Lorena Dini.

Screening and eligibility assessment: Sydney Odonkor, Ferdinand Koranteng

Quality assessment: Sydney Odonkor, Ferdinand Koranteng

Data extraction: Sydney Odonkor, Martin Appiah-Danquah

Data analysis: Sydney Odonkor

Writing – original draft: Sydney Odonkor.

Editing and revision of drafts: Lorena Dini

Approval of final version: all authors

## ETHICAL CONSIDERATIONS

This study was a systematic review and as such did not require any ethical clearance.

## ACKNOWLEDGEMENTS

Our sincere gratitude goes to the entire management of MScIH-Charité Universitätsmedizin Berlin especially Mrs. Francine Kyeremaa and Dr. Angel Phuti, for their immense support. We are also very grateful to Michael Zobi MD MScIH for his guidance and support.

## SUPPORTING INFORMATION

**S1 Table. Search strategies:** This is a description of our comprehensive list of search strings used to identify all studies in the various electronic databases

**S2 Table. Complete data extraction form:** This contains the complete data extracted from the final included articles in this review.

**S3 Table.** PRISMA Checklist

