## Supplementary material for "Do National Health Insurance Schemes Guarantee Financial Risk Protection in the drive towards Universal Health Coverage in West Africa? A Systematic Review of Observational Studies": Search strategies: This is a description of our comprehensive list of search strings used to identify all studies in the various electronic databases

**Table 1: Search Strategies**

| Database | Search number | Search String | Articles found |
|---|---|---|---|
| PubMed/Medline | #1 | “Insurance, Health”[Mesh] OR “Universal Health Insurance”[Mesh] OR “National Health Programs”[Mesh] OR “health insurance*”[tw] | 275,513 |
| | #2 | “Africa, Western”[Mesh] OR “west* Africa*”[tw] OR Benin[tw] OR “Burkina Faso”[tw] OR “Cape Verde”[tw] OR “Cabo Verde“[tw] OR “Cote d’Ivoire”[tw] OR “Ivory Coast”[tw] OR Gambia[tw] OR Ghana[tw] OR Guinea[tw] OR “Guinea-Bissau”[tw] OR Liberia[tw] OR Mali[tw] OR Niger[tw] OR Nigeria[tw] OR Senegal[tw] OR “Sierra Leone”[tw] OR Togo[tw] | 273,215 |
| | #3 | “Risk Sharing, Financial”[Mesh] OR “Poverty”[Mesh] OR “financial risk”[tw] OR “catastrophic health expenditure*”[tw] OR “catastrophic loss*”[tw] OR “financial loss*”[tw] OR impoverish*[tw] OR poverty*[tw] | 75,397 |
| | #4 | “Universal Health Care”[Mesh] OR UHC[tw] OR "universal health"[tw] OR "health for all*"[tw] | 10,604 |
| | #5 | #1 AND #2 AND (#3 OR #4), from 2005 - 2022 | **327** |
| Web of Science | #1 | ALL=("Health Insurance*") | 51,240 |
| | #2 | (ALL=("west* Africa*") OR ALL=(Benin) OR ALL=("Burkina Faso") OR ALL=(“Cape Verde”) OR ALL=(“Cabo Verde“) OR ALL=(“Cote d'Ivoire”) OR ALL=(“Ivory Coast”) OR ALL=(Gambia) OR ALL=(Ghana) OR ALL=(Guinea) OR ALL=(“Guinea-Bissau”) OR ALL=(Liberia) OR ALL=(Mali) OR ALL=(Niger) OR ALL=(Nigeria) OR ALL=(Senegal) OR ALL=(“Sierra Leone”) OR ALL=(Togo)) | 451,349 |
| | #3 | (ALL=("Financial risk") OR ALL=("catastrophic health expenditure*") OR ALL=("catastrophic loss*") OR ALL=("financial loss*") OR ALL=(impoverish*) OR ALL=(poverty)) | 116,671 |
| | #4 | (ALL=(“Universal health") OR ALL=(UHC) OR ALL=("health for all")) | 10,644 |
| | #5 | #1 AND #2 AND (#3 OR #4), from 2005 - 2022 | **293** |

| Database | Search number | Search String | Articles found |
|---|---|---|---|
| CINAHL via EBSCOhost | #1 | "Health Insurance*" | 400,602 |
| | #2 | "west* Africa*" OR Benin OR "Burkina Faso" OR “Cape Verde OR “Cabo Verde” OR “Cote d'Ivoire” OR “Ivory Coast” OR Gambia OR Ghana OR Guinea OR “Guinea-Bissau” OR Liberia OR Mali OR Niger OR Nigeria OR Senegal OR “Sierra Leone” OR Togo | 146,028 |
| | #3 | "Financial risk” OR "catastrophic health expenditure*" OR "catastrophic loss*" OR "financial loss*" OR impoverish* OR poverty | 640,030 |
| | #4 | “Universal health" OR UHC OR "health for all" | 51,896 |
| | #5 | #1 AND #2 AND (#3 OR #4), from 2005 - 2022 | **262** |
| Embase via Ovid | #1 | health insurance/ or health insurance*.mp. | 181,001 |
| | #2 | (West* Africa* or Benin or Burkina Faso or Cape Verde or Cabo Verde or Cote d'Ivoire or Ivory Coast or Gambia or Ghana or Guinea or Guinea-Bissau or Liberia or Mali or Niger or Nigeria or Senegal or Sierra Leone or Togo).mp. | 33,111 |
| | #3 | (Financial risk or catastrophic health expenditure or catastrophic loss or financial loss).mp. or poverty/ or poverty.mp. or impoverish*.mp. | 74,354 |
| | #4 | universal health care/ or universal health.mp. or UHC.mp. or health for all.mp. | 19,670 |
| | #5 | #1 AND #2 AND (#3 OR #4), from 2005 - 2022 | **385** |
| Google scholar | #1 | "Health insurance" AND "West Africa" AND "financial risk" OR "catastrophic health expenditure" OR "catastrophic loss" OR "financial loss" OR impoverish OR "universal health" | 5,184 |
| | Articles were ranked automatically based on relevance after which the first 500 articles were manually screened to find articles to be included at the titles and abstract screening stage. | | **12** |
